# Factor V is an immune inhibitor that is expressed at increased levels in leukocytes of patients with severe Covid-19

**DOI:** 10.1101/2021.01.14.21249801

**Authors:** Jun Wang, Prasanti Kotagiri, Paul A Lyons, Federica Mescia, Laura Bergamaschi, Lorinda Turner, Rafia S Al-Lamki, Michael D Morgan, Fernando J Calero-Nieto, Karsten Bach, Nicole Mende, Nicola K Wilson, Emily R Watts, Cambridge Institute of Therapeutic Immunology and Infectious Disease-National Institute of Health Research (CITIID-NIHR) COVID BioResource Collaboration, Patrick F Chinnery, Nathalie Kingston, Sofia Papadia, Kathleen Stirrups, Neil Walker, Ravindra K Gupta, Mark Toshner, Michael P Weekes, James A Nathan, Sarah R Walmsley, Willem H Ouwehand, Mary Kasanicki, Berthold Göttgens, John C Marioni, Kenneth GC Smith, Jordan S Pober, John R Bradley

## Abstract

Severe Covid-19 is associated with elevated plasma Factor V (FV) and increased risk of thromboembolism. We report that neutrophils, T regulatory cells (Tregs), and monocytes from patients with severe Covid-19 express FV, and expression correlates with T cell lymphopenia. *In vitro* full length FV, but not FV activated by thrombin cleavage, suppresses T cell proliferation. Increased and prolonged FV expression by cells of the innate and adaptive immune systems may contribute to lymphopenia in severe Covid-19. Activation by thrombin destroys the immunosuppressive properties of FV. Anticoagulation in Covid-19 patients may have the unintended consequence of suppressing the adaptive immune system.

## Introduction

Dysregulation of both the immune^1^ and coagulation system^2^ occurs in Covid-19 infection. Immunological responses include T cell lymphopenia, which we have found can persist for months after the initial illness. Coagulopathy is an important cause of morbidity and mortality in patients with Covid-19, and a marked increase in circulating FV activity has been reported in patients with severe Covid-19, associated with increased risk of thromboembolism^3^.

Production of FV by T cells^4^ and monocytes^5^ has been previously reported. We report that circulating leukocytes are a source of FV in patients with Covid-19, and propose that neutrophil, monocyte and Treg derived Factor V may be an important determinant of the dysregulated immune response to SARS-CoV-2.

## Results

### FV is produced by circulating blood cells and increased in hospitalised patients with Covid-19

Analysis of the transcriptome of peripheral blood cells from healthy controls and patients with Covid-19 express FV, and expression is increased in patients with more severe disease for at least 60 days after onset of symptoms (Figure 1A). Analysis of the Blueprint (https://www.blueprint-epigenome.eu/) database of haematopoietic genome-scale datasets shows the highest level of FV expression in neutrophils, eosinophils, Tregs and monocytes (Supplementary Figure 1). To investigate whether Tregs and monocytes express FV in patients with Covid-19 we performed scRNAseq on peripheral blood mononuclear cells (PBMCs) derived from healthy controls and patients with Covid19. Tregs (CD4+, FoxP3+) had the highest expression of FV, with expression also detected in monocytes and lower levels in other CD4 cell subsets (Figure 1B). However, expression in PBMCs did not appear to correlate with disease severity. Expression of FV in Tregs from healthy controls was confirmed by RT-PCR and immunoblotting (Supplementary Figure 2).

**Figure 1.**
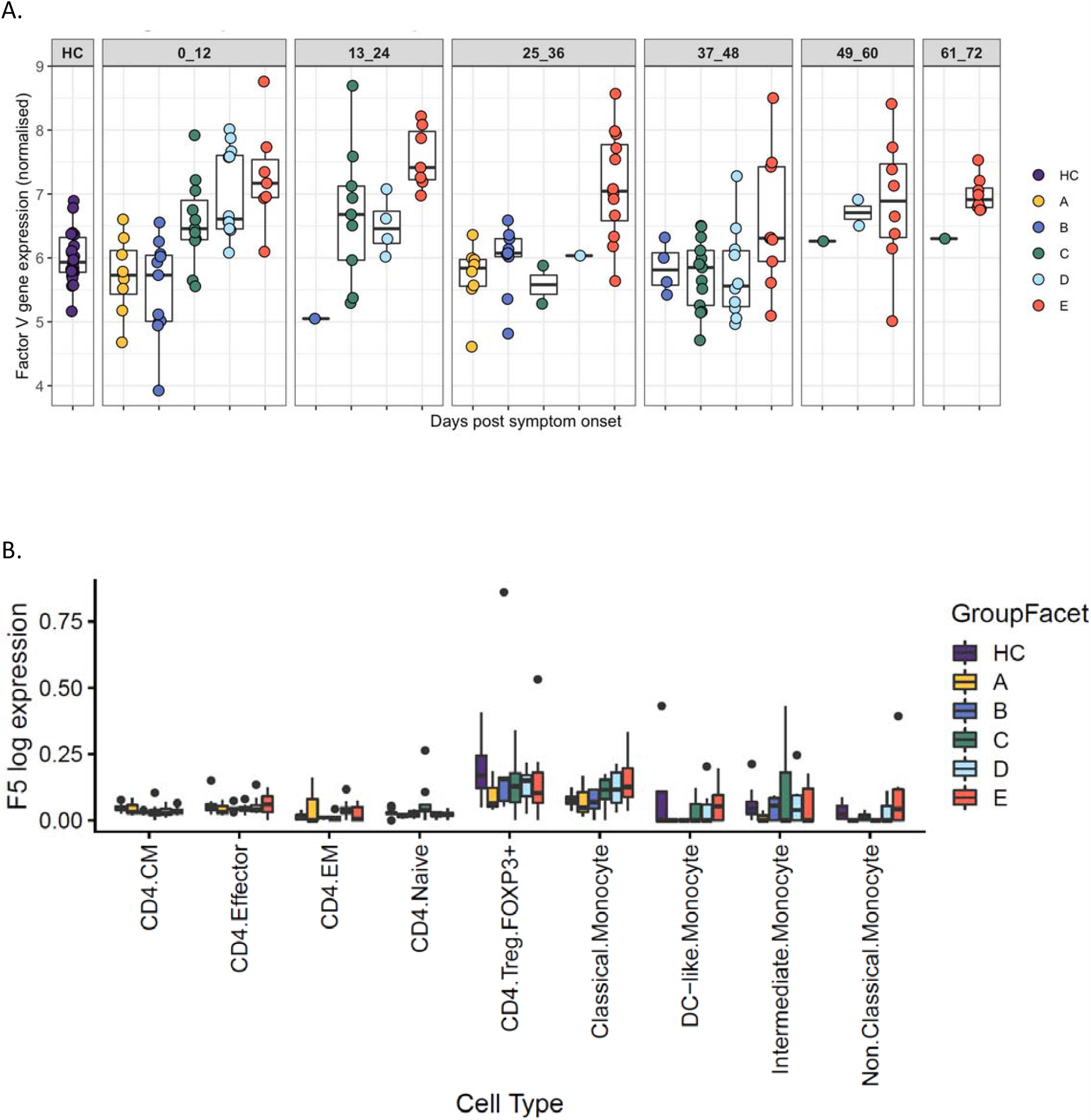
FV is produced by circulating blood cells and increased in hospitalised patients with Covid-19. A. Peripheral blood cells from healthy controls, healthcare workers and patients with Covid-19 express FV. Individuals represented by dots are grouped into 12 day time periods after onset of symptoms or a positive swab in asymptomatic healthcare workers (HCW). HC, healthy controls; A, HCW screening asymptomatic; B, HCW screening symptomatic; C, hospitalised mild disease; D, hospitalised requiring oxygen; E, hospitalised, intensive care. 0 to 12 days D vs HC p = 0.0007; E vs HC p = 0.0002. 13 to 24 days E vs HC p = 0.00001. 25 – 36 days E vs HC p = 0.00003. 49 to 60 days E vs HC p = 0.013. 61 to 72 days E vs HC p = 0.00003. B. scRNAseq of PBMCs derived from HC, HCW and patients with Covid-19 showed the highest expression of FV in CD4+, FoxP3+ Tregs, with expression also detected in monocytes and at lower levels in other CD4 cell subsets. FV expression was increased in CD4+ Naïve vs CD4+ FOXP3 Tregs in healthy controls (p= 8.33×10^−4^), and in severe Covid-19 vs healthy controls in classical monocytes but not other cell subsets (p = 0.016).

### Neutrophils are a source of increased FV expression in patients with severe Covid-19

Weighted gene co-expression network analysis was performed to create distinct modules comprising of non-overlapping co-expressed genes. The module containing FV also contained genes strongly expressed in neutrophils (Figure 2A, Supplementary Figure 3). In addition, FV was a “hub gene” meaning its expression closely mirrored that of the module eigengene, which is a single expression profile summarising all genes within the module. This FV module eigengene expression correlated with severity of Covid-19 (Figure 2B). FV module expression is similar to healthy controls in asymptomatic or mildly symptomatic individuals in the community. In hospitalised patients with mild disease FV module expression is elevated at presentation, but declines as patients recover. In patients with severe disease FV module expression is elevated at presentation and increased levels persists for several weeks. Analysis of peripheral blood cells for expression of other coagulation factors showed expression of Factor XIIIa and low levels of Factor XII but not other coagulation factors (data not shown).

**Figure 2.**
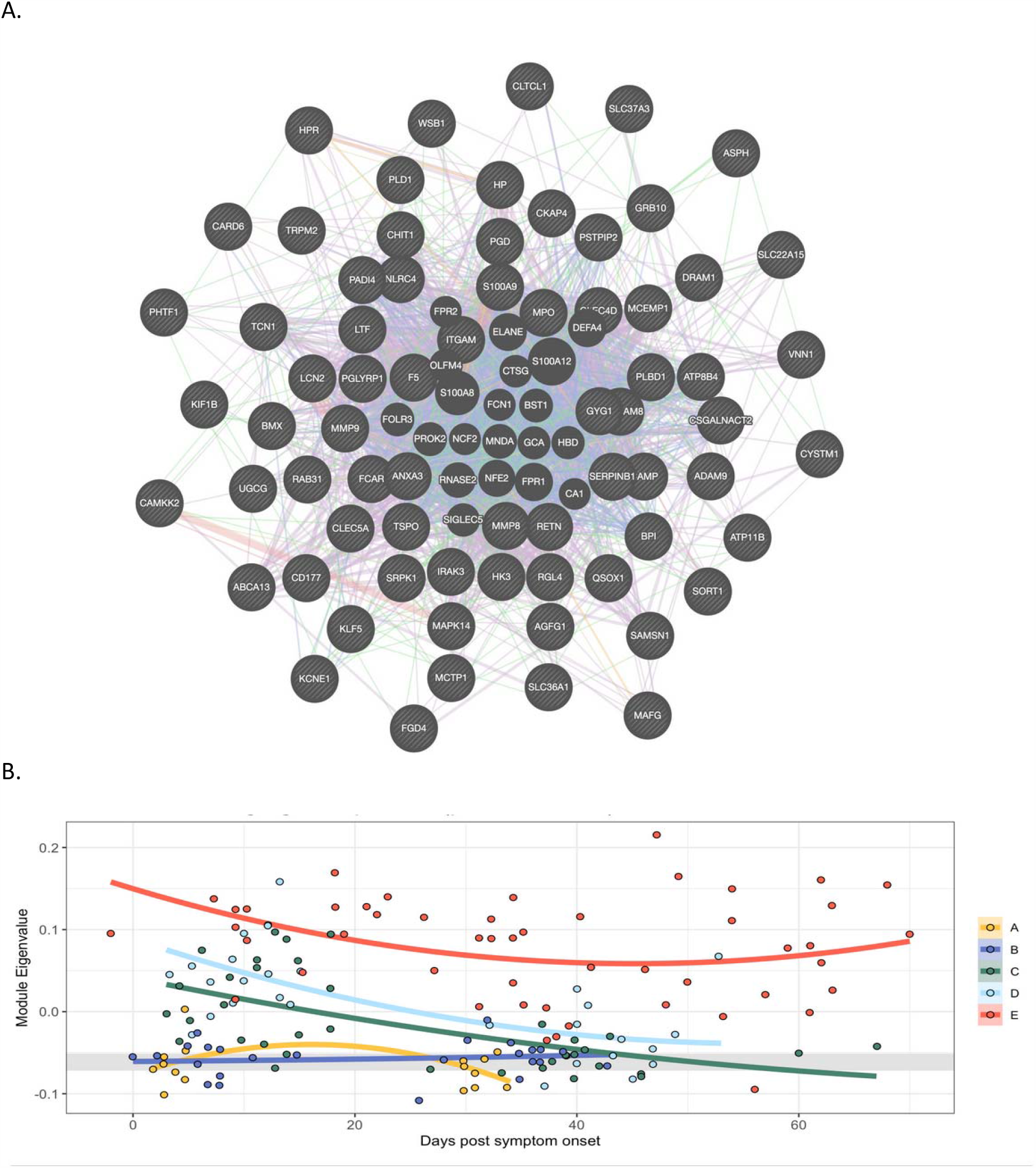
In peripheral blood cells FV is co-expressed with a module of genes expressed in neutrophils, and expression of the module correlates with severity of Covid-19. A. Weighted gene co-expression network analysis identified a module containing group of genes co-expressed with FV. In this module FV is a hub gene and its expression correlates strongly with genes expressed in neutrophils. The lines are colour coded to show relationships. Purple: Gene known to be co-expressed in existing gene databases. Orange: Predicted functional relationships between genes. Pink: Proteins known to be linked. Turquoise: Genes present in a shared annotated pathway. Blue: Genes expressed in the same tissue. B. Mixed-effects model with quadratic time trend showing the longitudinal expression of the FV module over time, grouped by severity. Grey band indicates the interquartile range of HCs. A significant effect of time versus severity group interaction term (p = 0.0047) indicates that disease severity has a significant effect on longitudinal expression. A, HCW screening asymptomatic; B, HCW screening symptomatic; C, hospitalised mild disease; D, hospitalised requiring oxygen; E, hospitalised, intensive care.

To determine whether increased peripheral blood cell FV mRNA levels correlate with FV protein expression we assayed plasma FV levels and performed liquid chromatography – mass spectrometry on neutrophil lysates from healthy controls and patients with severe Covid-19. There was a modest but statistically significant correlation between FV gene expression and FV protein expression (Figure 3A; p = 0.023, R = 0.17). Proteomic analysis showed significantly higher levels of FV in neutrophil lysates from patients with severe Covid-19 compared to healthy controls (Figure 3B).

**Figure 3.**
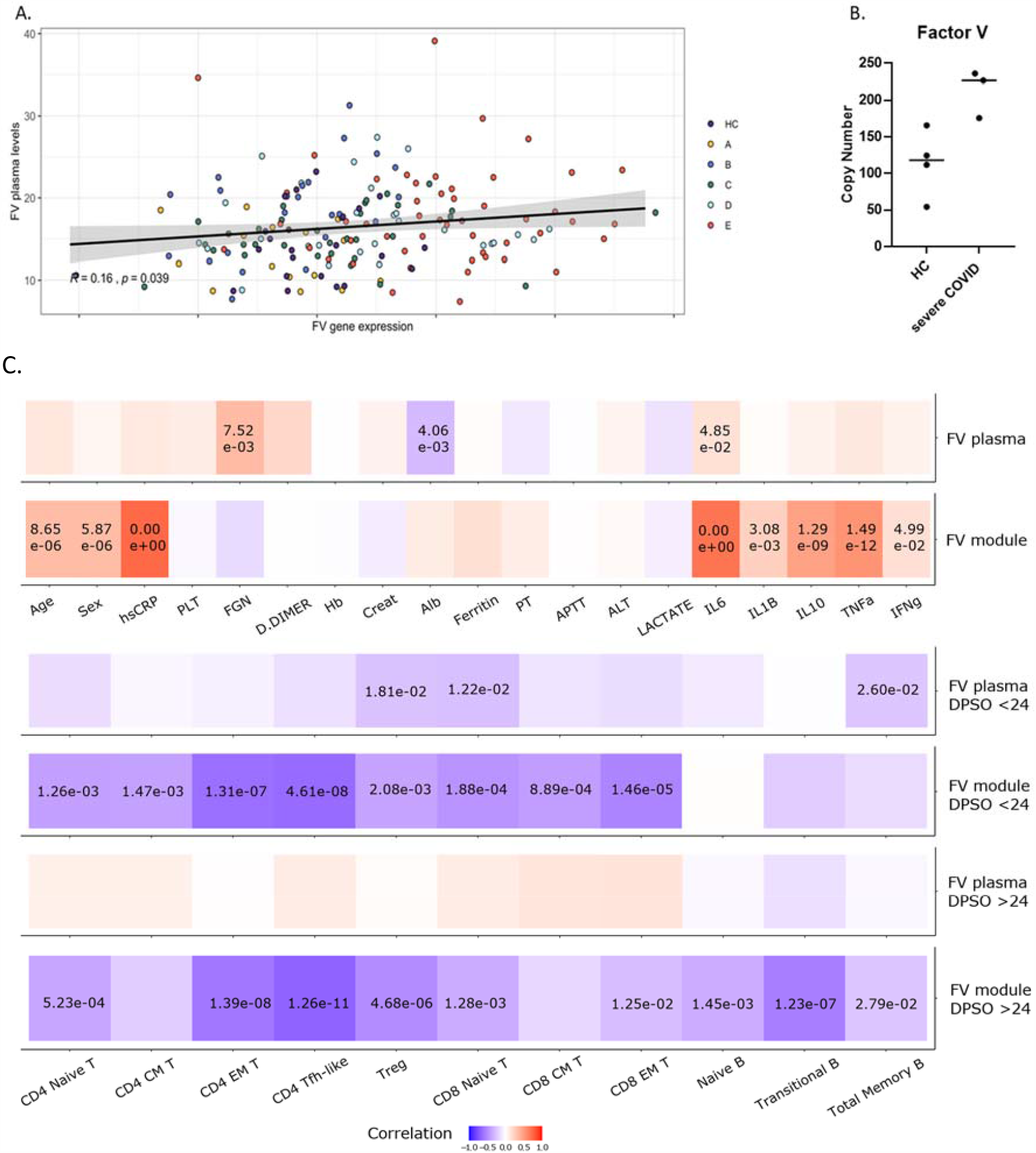
FV expression correlates with parameters of disease severity and lymphopenia. A. Correlation of plasma FV levels and peripheral blood cell FV gene expression in healthy controls and patients with Covid-19 (r=0.16, p = 0.039). B. LC-MS measurement of FV in protein lysates of neutrophil extracts from healthy controls and patients with severe Covid-19. FV levels were significantly higher in neutrophil lysates from patients with severe Covid-19 compared to healthy (p = 0.025). C. Correlation of plasma FV levels and FV module gene expression with predictors of disease severity and plasma protein levels (i-ii); or with B and T cell counts (iii-vi). FV plasma levels correlate with fibrinogen and IL6 (i), whereas FV module gene expression correlated with predictors of disease severity (age, male gender, CRP) and increased plasma levels of IL6, IL10 and TNF (ii). There was very little correlation between plasma factor V levels and T and B cell counts during the first 24 days post symptom onset (DPSO, iii) or after 24 days from the onset of symptoms (v). In contrast, FV module gene expression correlates with suppression of T cell counts during the first 24 days post symptom onset (iv), and T and B cell counts after 24 days from the onset of symptoms (vi). p values are shown where significant.

### FV expression correlates with parameters of disease severity and lymphopenia

FV module gene expression correlates with suppression of T cell counts during the first 24 days after symptom onset (Figure 3C.iv), and T and B cell counts after 24 days from the onset of symptoms (Figure 3C.vi). In contrast, there was very little correlation between plasma factor V levels and T and B cell counts during the first 24 days after symptom onset (Figure 3C.iii) or after 24 days from the onset of symptoms (Figure 3C.v). FV module gene expression correlated with predictors of disease severity (age, male gender, CRP) and increased plasma levels of IL6, IL10 and TNF, but not clinical parameters including laboratory measurements of haemostasis (Figure 3C.ii), whereas FV plasma levels correlate with laboratory measures of haemostasis (Figure 3C.i).

### FV suppresses T cell proliferation

The high level of expression of FV in Tregs led us to determine if FV suppresses *in vitro* proliferation of T cells. To examine this, CFSE-labelled conventional CD4+ T cells (Tcons) from healthy donors were stimulated *in vitro* and proliferation was assessed by flow cytometry analysis of dye dilution. FV but not FV activated by thrombin (FVa) suppressed proliferation of Tcons in a concentration dependent manner (Figure 4). To confirm that the suppressive effect was mediated by full length uncleaved FV we generated recombinant proteins from three FV constructs: (1) FV(738-1573)-6His (the B domain of FV); (2) FV-6His (full length Factor V); (3) Factor V R709A, R1018A, R1545A-6His (full length FV with Arginine thrombin cleavage sites mutated). Full length recombinant FV, but not a recombinant B domain, inhibited Tcon proliferation, and this effect was prevented by thrombin, and enhanced by the thrombin inhibitor hirudin (Figure 5A). Thrombin did not increase proliferation in the absence of FV. A cleavage resistant recombinant FV was a potent inhibitor of Tcon proliferation. Recombinant full length FV and a cleavage resistant recombinant FV also suppress CD8+ T cell proliferation, but not B cell proliferation (Figure 5B).

**Figure 4.**
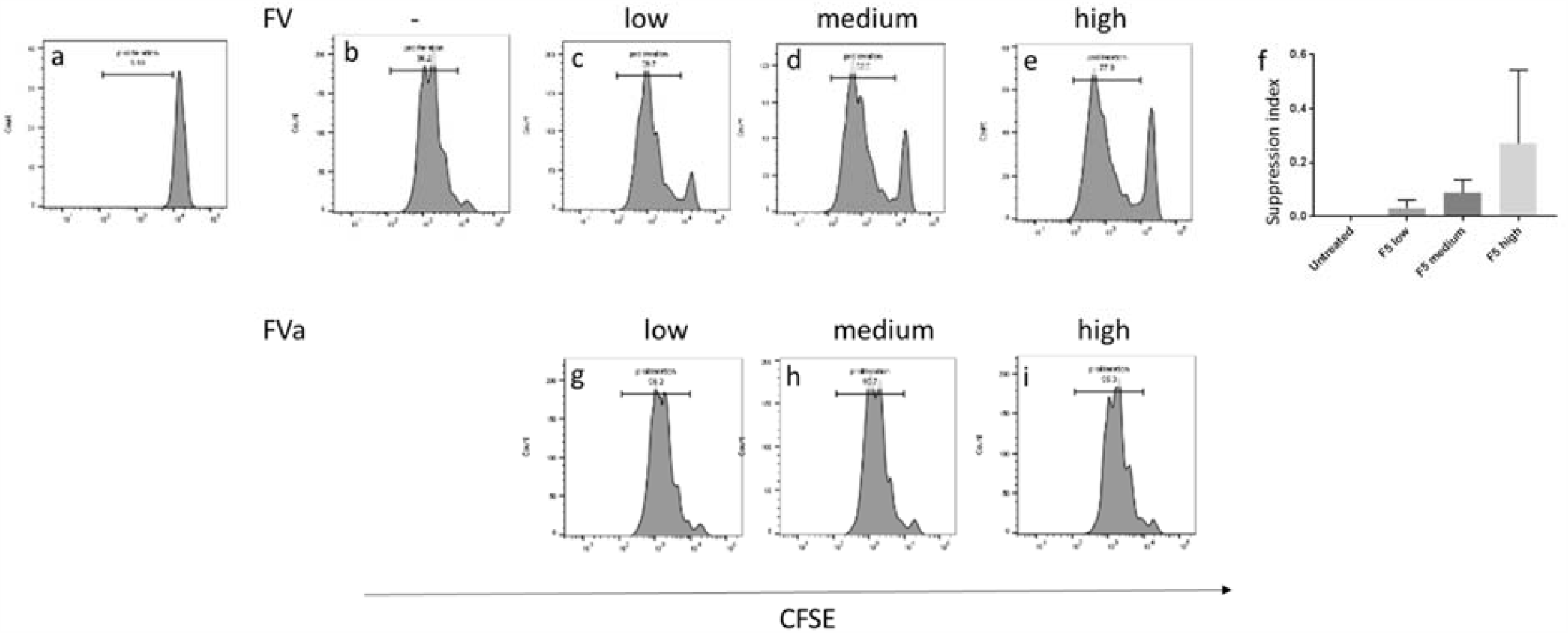
FV suppresses T cell proliferation. CD4+T conventional cells (Tcon) were labelled with carboxyfluorescein diacetate succinimidyl ester (CFSE) (a), and stimulated with Dynabeads T cell activator, proliferation was shown as diluting the dye and reducing fluorescence intensity (b). Proliferation is inhibited in a concentration dependent manner by addition of native Factor V (panels c – e), but not by activated factor V, FVa (g – i). Data are representative of 10 healthy donors (f), FV at medium concentration suppressed T cell proliferation by 3 to 17 percent (p=0.0003). Medium concentration is close to the physiological plasma level at 20 nM, low concentration is 4 nM, high concentration is 100 nM.

**Figure 5.**
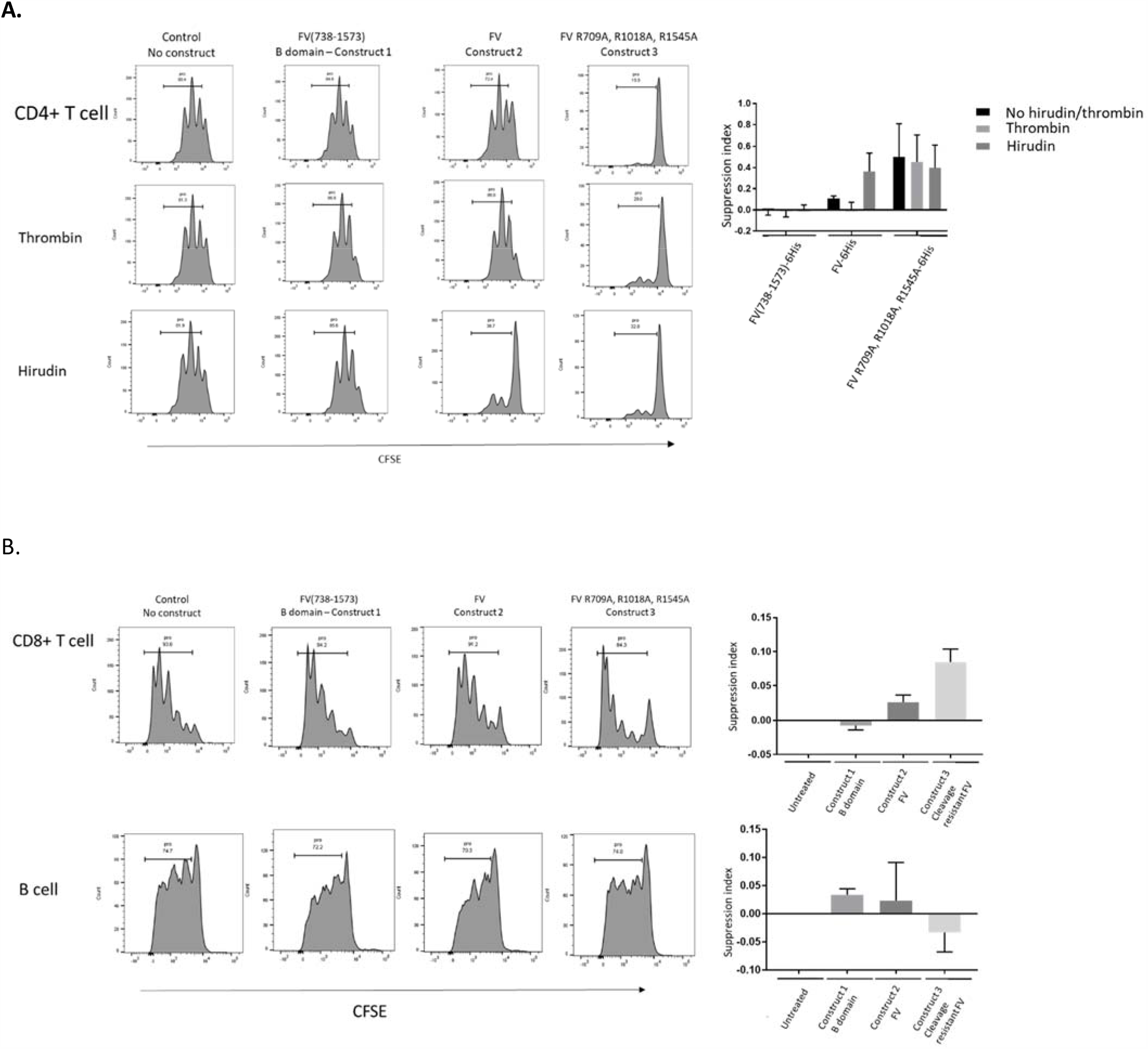
Full length, but not thrombin cleaved FV, suppresses T but not B cell proliferation. A. Tcon proliferation was not inhibited by recombinant FV B domain (Construct 1), and thrombin and hirudin had no effect on their own and in combination with construct 1. Recombinant full length Factor V (construct 2) inhibited Tcon proliferation similar to native plasma derived Factor V, and its effect was prevented by thrombin (p=0.036), while the effect of inhibition by mutated Factor V (construct 3) was not prevented by thrombin (p=0.25). B. CD8+ T cell proliferation was inhibited similarly by construct 2 (p=0.009) and construct 3 (p=0.004), while B cell proliferation was not inhibited by any of the constructs (p=0.1; p=0.3; p=0.6). Pro: prolifertation.

## Discussion

Covid-19 is associated with an increased risk of venous thromboembolism (VTE), particularly in patients with severe illness. Abnormalities of the coagulation system are likely to contribute to the increased risk, and the most consistent laboratory finding is an increased D-dimer level, with some reports of prolongation of the prothrombin time and thrombin time, and shortened activated partial thromboplastin time^6,7^. FV levels are increased in Covid-19^8^, and marked increases in FV activity have been associated with VTE in some patients with severe Covid-19^3^.

Our data show that circulating neutrophils, monocytes and Tregs are a source of FV in patients with severe Covid-19, and also identify Tregs as a source of FV in healthy blood donors. Furthermore FV decreases T cell proliferation *in vitro*. This suppression of T cell responses is lost if FV is activated by thrombin, and potentiated by a recombinant FV in which thrombin cleavage sites are mutated to prevent activation. In patients with Covid-19 increased circulating blood cell FV expression is associated with suppression of T cell counts, and both increased FV expression and suppressed T cell subsets can persist for several months after infection with SARS-CoV-2.

There was a modest but statistically significant correlation between FV gene expression in circulating blood cells and plasma FV levels, but the key role of leukocyte derived FV in regulating the immune response to SARS-CoV-2 is likely to occur following migration of leukocytes into the tissues, including secondary lymphoid organs from which circulating FV is normally excluded. Broncho-alveolar lavage fluid from patients with severe Covid-19 has shown a predominance of neutrophils, monocytes/macrophages, and eosinophils with few lymphocytes, and in particular CD4 and CD8 T cell lymphopenia, but an increase in the proportion of Tregs^9^. Furthermore lymphocytes were either absent or sparse in areas of SARs-CoV-2 pneumonia infiltrated by macrophages or neutrophils, or containing neutrophil extracellular traps (NETs) composed of neutrophil DNA, histones and granule-derived enzymes^10,11^. Thus leukocyte derived FV may suppress local T cell proliferation at sites of infection. In support of this plasma FV levels correlated with biomarkers of haemostasis, whereas T cell counts correlated with FV module gene expression in circulating leukocytes. The liver may be the predominant source of circulating FV, and it is noteworthy that the liver displays adaptive immune tolerance^12^.

Guidelines recommend unfractionated heparin (UFH) or low molecular weight heparin (LMWH) as prophylaxis against VTE in Covid-19^13^. Heparin binds to and activates anti-thrombin^14^. LMWHs retain some anti-thrombin activity, but most of their activity is thought to result from inhibition of Factor Xa^15^. Alternatives for VTE prophylaxis include warfarin and direct oral anticoagulants (DOACs). Warfarin depletes the reduced form of vitamin K that acts as a cofactor for gamma carboxylation of prothrombin, VII, IX and X, rendering them inactive^16^. DOACs inhibit factor Xa or thrombin. The effect of anticoagulants on circulating factor V levels is unknown, but by inhibiting thrombin heparin reduces factor V activation, and could potentiate the T cell suppressive effect of FV. In support of this there is evidence that heparin can directly suppress immune, and in particular T cell, responses^17^. Prophylaxis against VTE is an essential for the management of hospitalised patients with Covid-19. Increased FV expression by cells of the innate and adaptive immune systems may explain the lymphopenia seen in patients with severe Covid19. Heparin may potentiate suppression of the adaptive immune response by reducing FV activation.

## Methods

### Materials

Rabbit anti-human FoxP3 (Cell signalling technology,Hitchin, UK). Mouse anti-human FV (Cambridge Biosciences, Cambridge, UK). B cell proliferation kit (R&D system, Abingdon, UK). BD CellFix (BD Biosciences, Oxford, UK). Proteinase inhibitor cocktail (Roche Diagnostics Ltd, West Sussex, UK). Native human FV, FVa, Thrombin, Dynabeads Human T cell activator, PCR kits, CYBRgreen kit and CFSE kits (Thermofisher Scientific, Loughborough, UK). CD4+CD25 + CD127-T reg, CD8 T cell and B cell isolation kit (Miltenyi Biotec, Surrey, UK). Assay of FV plasma levels was performed using a human Factor V ELISA Kit (ab137976, Abcam, Cambridge, UK). Unless otherwise indicated, all reagents were from Sigma-Aldrich Company Ltd (Dorset, UK).

### Healthy volunteers and patients

Healthy donor blood samples were obtained from the NIHR Cambridge BioResource and leukapheresis samples from the National Health Service Blood and Transfusion services (NHSBT, Cambridge) with written informed consent of donors and approval of the National Research Ethics Committee and Health Research Authority. Healthcare workers and patients with Covid-19 confirmed by Nucleic acid amplification testing^18^ of nasopharyngeal swabs for SARS-CoV-2 were consented to the NIHR Covid-19 cohort of the NIHR BioResource (https://cambridgebrc.nihr.ac.uk/bioresourcecovid19/) with approval of the National Research Ethics Committee and Health Research Authority. These included patients presenting to Cambridge University Hospitals and Royal Papworth Hospital, together with asymptomatic or symptomatic healthcare workers (HCWs) undergoing routine screening. Samples were also collected from seronegative concurrent controls recruited. Hospitalised patients underwent a venous thromboembolism risk assessment and received prophylactic dalteparin if there was low bleeding risk. From 1 May 2020 patients were discharged with 2 weeks of prophylactic dalteparin following a risk assessment.

For neutrophil proteomic studies peripheral venous blood was taken from healthy volunteers (age 25 – 60, one female) with written informed and approval of the University of Edinburgh Centre for Inflammation Research Blood Resource Management Committee. The collection of peripheral venous blood from patients (age 41 – 56, one female) with Covid-19 was approved by Scotland A Research Ethics Committee. Patients receiving ventilation in intensive care at the Royal Infirmary of Edinburgh were recruited between April and August 2020, with informed consent obtained by proxy.

### Peripheral blood cell isolation

PBMCs were isolated from blood by polysucrose density gradient centrifugation (Ficoll-Paque, GE Healthcare Life Sciences, UK). Cells were grown in RPMI media supplemented with 10% human AB serum.

Neutrophils were isolated from blood by dextran sedimentation and discontinuous Percoll gradients. Analysis of neutrophil lysates was performed using liquid chromatography mass spectrometry (LC-MS)-based proteomics

### T cell *in vitro* expansion assay

Tcons were isolated with the above-mentioned kit. Cells were stained with CFSE following the manufacturer’s instruction. 5 × 10^4^ labelled T cons were mixed with Dynabeads Human T cell activator CD3/CD28 at cell-to-beads ratio of 2 to 1. Cells were collected and analysed using a BD Fortessa flow cytometer after 4 or 5 days. Suppression index was calculated as the ratio between decreased percentage of proliferation and the total percentage of proliferation of the cells.

Flow cytometry data were analysed using FlowJo (Tree Star, USA). Graphs and statistics were generated using GraphPad Prism software. Results were presented as mean ± s.e.m.as indicated. Differences between two groups were compared using two-tailed student’s t-test.

### Whole blood bulk mRNA-Seq

#### Library preparation and RNA-Seq processing

RNA was quantified using RNA HS assay on the Qubit, and libraries prepared using the SMARTer^®^ Stranded Total RNA-Seq it v2 - Pico Input Mammalian kit (Takara) with 10ng of RNA as starting input. Library quality and quantity were validated by capillary electrophoresis on an Agilent 4200 TapeStation. Libraries were pooled at equimolar concentrations, and paired-end sequenced (75bp) across 4 lanes of a Hiseq4000 instrument (Illumina) to achieve 10 million reads per samples.

#### Reads mapping and quantification

The quality of raw reads was assessed using FastQC (http://www.bioinformatics.babraham.ac.uk/projects/fastqc/). SMARTer adaptors were trimmed, along with sequencing calls with a Phred score below 24 using Trim_galore v.0.6.4 (http://www.bioinformatics.babraham.ac.uk/projects/trim_galore/.) Residual rRNA reads were depleted in silico using BBSplit (https://github.com/BioInfoTools/BBMap/blob/master/sh/bbsplit.sh). Alignment was performed using HISAT2 v.2.1.0^19^ against the GRCh38 genome build achieving a more than 95% alignment rate. A count matrix was generated in R using featureCounts (Rsubreads - packages) and converted into a DGEList (EdgeR package), for downstream analysis.

#### Downstream Analysis

Downstream analysis was performed in R. Counts were filtered using filterByExpr (EdgeR package) with a gene count threshold of 10CPM and the minimum number of samples set as the size of the smallest disease group. Library counts were normalised using calcNormFactors (EdgeR package) using the method ‘weighted trimmed mean of M-values’. The function ‘voom’ (limma package) was applied to the data to estimate the mean-variance relationship, allowing adjustment for heteroscedasticity.

#### Weighted gene co-expression network analysis

The weighted gene co-expression network analysis (WGCNA) package in R overcomes the problem of multiple testing by grouping co-correlated genes into modules and relating them to clinic traits. Modules are not comprised of a priori defined gene sets but rather are generated from unsupervised clustering. The eigengene of the module is then correlated with the sample traits and significance determined. A signed adjacency matrix was generated, and a soft thresholding power chosen to impose approximate scale-free topology. Modules identified from the topological overlap matrix had a specified minimum module size of 30. Significance of correlation between a clinical trait and a modular eigengene was assessed using linear regression with Bonferroni adjustment to correct for multiple testing. Modules were annotated using Enrichr and Genemania. Genes with high connectivity termed “hub genes” were identified based on a module membership of 0.8 or above and were selected to have a correlation with the trait of interest ≥0.8.

#### Correlation

The relationships between multiple features were quantified using Pearson’s correlation (Hmisc package) and visualized with corrplot.

#### Mixed Effects model

Longitudinal mixed modelling of factor V module eigenvalue changes over time (*y*_*ij*_) was conducted using the nlme package in R, including time (*t*_*ij*_) with a quadratic trend and disease severity (*X*_*j*_) as fixed effects, and sampled individuals as random effects (*u*_*j*_).

### scRNAseq of PBMCs

#### Sample processing and antibody staining

Pools of purified PBMCs were generated by combining 0.5 million live cells from four individuals and viability assessed by counting in an improved Neubauer chamber using Trypan blue. Half a million viable cells were incubated with 2.5 μl of Human TruStain FcX™ Fc Blocking Reagent (BioLegend 422302), followed by 25 μl TotalSeq-C™ antibody cocktail (BioLegend 99813).

#### 10X Genomics droplet single-cell RNA-sequencing

50,000 live cells for each pool were processed using Single Cell VDJ 5’ version 1.1 (1000020) together with Single Cell 5’ Feature Barcode library kit (1000080), Single Cell V(D)J Enrichment Kit, Human B Cells (1000016) and Single Cell V(D)J Enrichment Kit, Human T Cells (1000005) from 10X Genomics following manufacturer’s recommendations. The samples were subjected to 12 cycles of cDNA amplification and 8 cycles for the protein library construction. The rest of libraries were processed as indicated by the manufacturer. Libraries were pooled per sample using the ratio 9:2.4:1:0.6 for gene expression, feature barcoding, TCR enriched and BCR enriched libraries. Samples were sequenced on an Illumina NovaSeq6000 sequencer machine using S1 flowcells.

#### Single-cell RNA-sequencing processing, demultiplexing and quality control

Multiplexed 10X scRNA-seq GEX libraries were aligned to the human genome, reads deduplicated, and UMIs quantified using Cellranger v4.0 utilising the hg38 genome reference sequence. Gene expression count matrices of genes by droplets were generated separately for each multiplex pool of donors^20^, demultiplexing and single cell genotypes derived using CellSNP (https://github.com/single-cell-genetics/cellSNP) were assigned to individual donors and quality control performed as previously described^21,22^. Doublets were identified using the “doubletCells” function in *scran* based on highly variable genes (HVGs). “Cluster walktrap” (https://arxiv.org/abs/physics/0512106) was used on the shared nearest-neighbour (SNN)-graph that was computed on HVGs in principal component space to form highly resolved clusters per sample. Doublets were removed prior to any downstream analyses.

#### Single-cell RNA-sequencing clustering and annotation

Highly variable genes (HVG) were defined across all single-cells and assigned to a donor individual. Single-cell clusters were computed by first constructing a k-nearest neighbour graph (k=20), and broadly grouped into discrete clusters based on the Walktrap community detection algorithm, using the kNN-graph as input. Each cluster was annotated into one of 7 broad categories based on the expression of canonical marker genes for each: CD4 T cell, CD8 Tcell, NK cell, Monocyte, Plasma cell, B cell and dendritic cell. HVGs, PCA, k-NN graph and clusters were re-computed for subsets of cells in each category, and sub clusters annotated based on marker genes.

Differential gene expression between clinical severity groups in Classical Monocytes, and between CD4+ Naïve and CD4+ FOXP3 Tregs in healthy controls, used the mean log normalized expression across single-cells from each individual. Mean expression values were compared in a linear model, p-values are reported from a 2-sided t-test adjusted for multiple testing using a Bonferroni correction (m=9 tests).

### RT-PCR

250 ng total RNA was amplified with FV forward (5′-ACCACAATCTACCATTTCAGGACTT −3′); FV reverse (5′-CGCCTCTGCTCACGAGTTAT −3′) and Foxp3 forward (5′-GCTGCAGCTCTCAACGGT −3′); Foxp3 reverse (5′-GGCAAACATGCGTGTGAAC −3′) using RT-PCR system following manufacture’s instruction ; the PCR product was visualized by 1% low melting temperature agarose gel in Tris-Acetate-EDTA buffer.

### Immunoblotting

Equal number of T regs and the CD4 enriched fraction isolated from the kit described above were collected and lysates prepared for SDS polyacrylamide gel electrophoresis and immunoblotting as previously described^23^. Signals were detected by enhanced chemiluminesence using ECL (Thermo Scientific, Paisley, UK). Images were collected and analysed using Image Lab (Bio-Rad, Hemel Hempstead, UK).

### Expression of FV constructs

Three constructs comprising (1) FV B domain ((aa710-1545), (2) full length FV (aa 1 − 2224) and (3) [R709A, R1018A, R1545A]FV aa1-2224 were sub-cloned into a proprietary vector for the HEK293-6E system. All sequences contained a C-terminal 6His tag to facilitate purification. Cells were transfected at a 500 ml scale for each construct, media harvested 5-6 days after transfection and protein purified using a combination of Ni affinity and size exclusion chromatography and if required ion exchange. Purified proteins were analysed by reducing and non-reducing SDS-PAGE, A280 to determine concentration, size exclusion and mass spectrometry to confirm identity.

## Data Availability

The NIHR BioResource makes data available to researchers through a Data Access Application process described at:
https://bioresource.nihr.ac.uk/using-our-bioresource/academic-and-clinical-researchers/apply-for-bioresource-data/

---

**Supplementary Figure 1.**
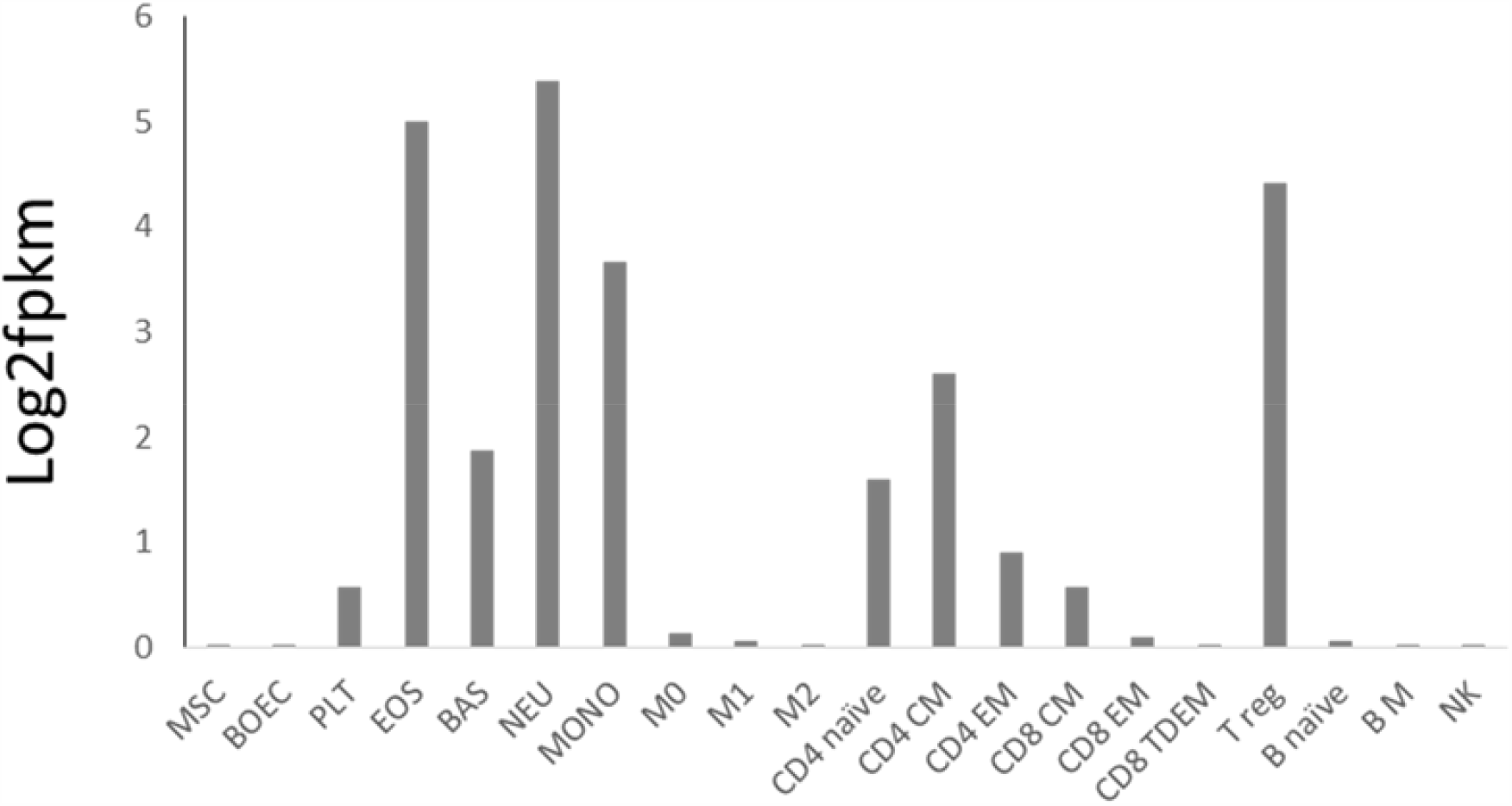
Analysis of the Blueprint (https://www.blueprint-epigenome.eu/) database identified FV expression in peripheral blood cell subsets in healthy individuals, with the highest expression in neutrophils, eosinophils, monocytes and Tregs derived from healthy individuals. fpkm - fragments per kilobase of transcript per million mapped reads. MSC: mesenchymal stem cells, BOEC: blood outgrowth endothelial cells, PLT: platelet, EOS: eosinophil, BAS: basophil, NEU: neutrophil, MONO: monocyte, M0, M1 and M2: macrophages, CM: central memory, EM: effector memory, TDEM: terminally differentiated effector memory, T reg: regulatory T cells, B M: B memory NK: natural killer.

**Supplementary Figure 2.**
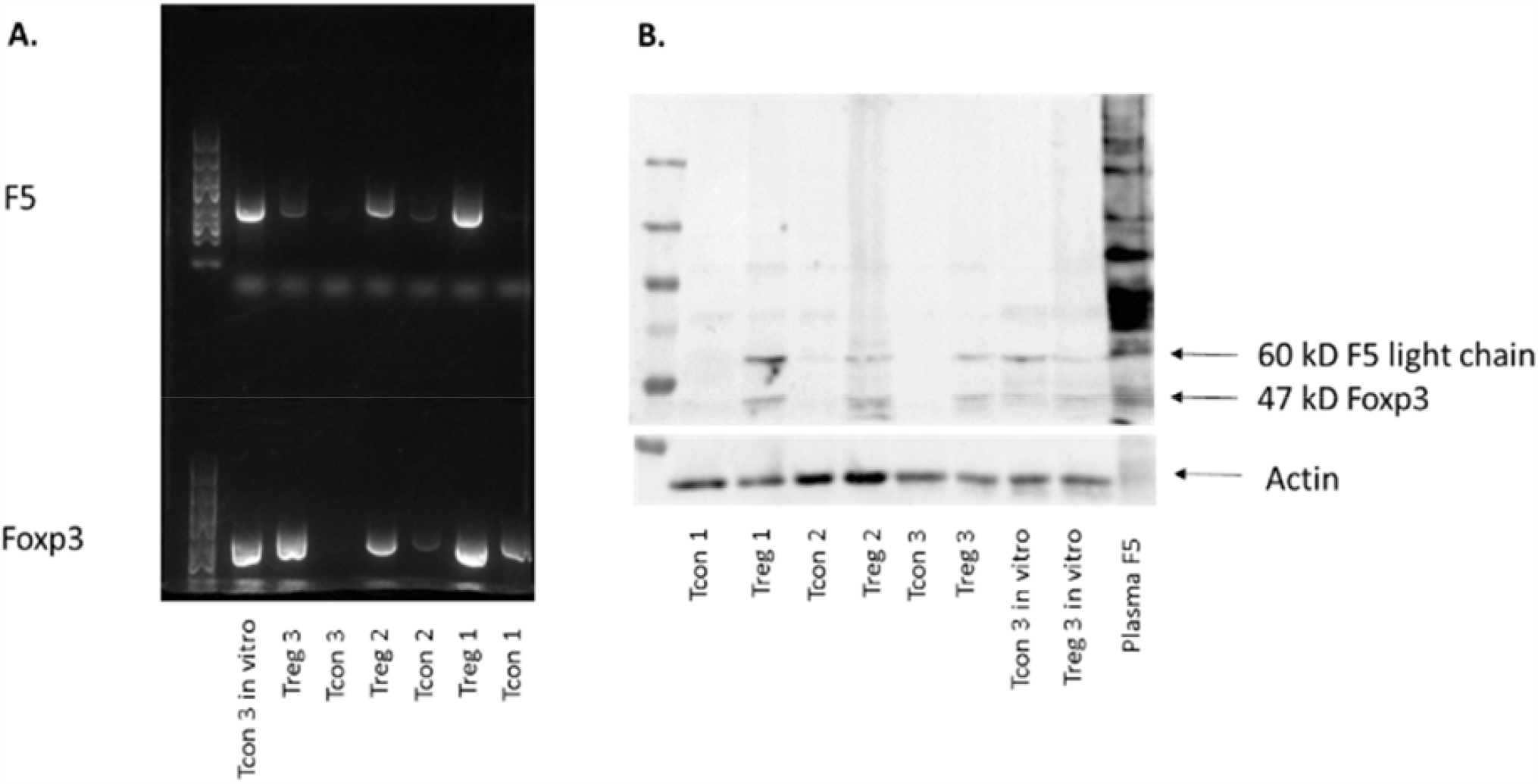
Expression of FV in Tregs, and also *in vitro* expanded T cells, was confirmed by Rt-PCR (A), and immunoblotting (B).

**Supplementary Figure 3.**
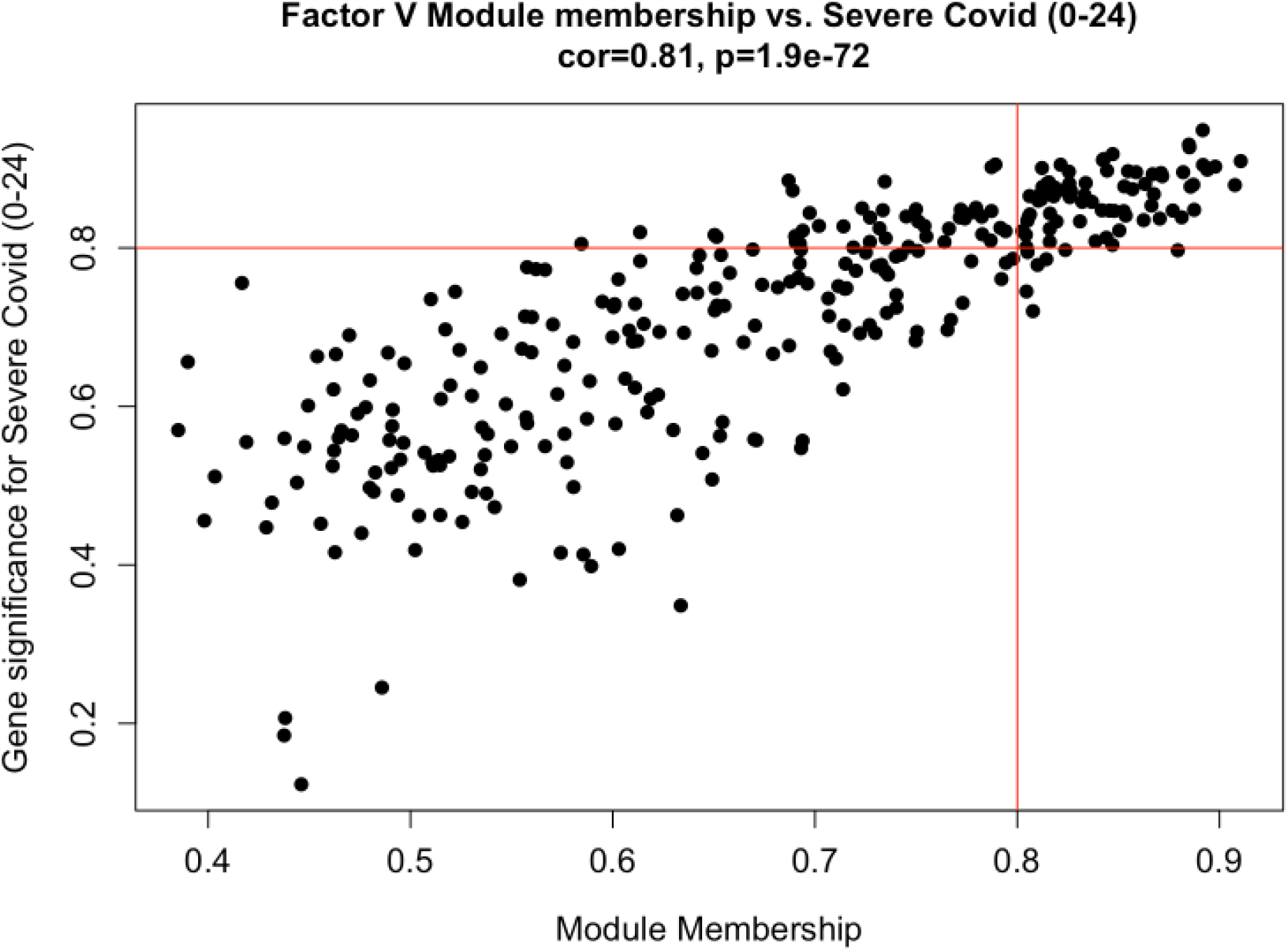
Factor V module membership vs gene significance for severe Covid-19. Genes comprising the Factor V module are graphed. The x axis represents the correlation of a given gene’s expression with the module eigenvalue. The higher the correlation, the more representative the module expression is of the gene. The Y axis represents the correlation between a gene’s expression and disease status (HC versus severe Covid-19 at 0-12 days post symptom onset). The higher the correlation, the more able a given gene can distinguish severe Covid-19 from health. The red lines demarcate the “hub genes” which represent genes that strongly model the module and disease status. The genes illustrated in Figure 2A are: KIF1B; PGD; PADI4; SORT1; PHTF1; SLC22A15; S100A9; F5; QSOX1; CHIT1; MTARC1; NLRC4; AGFG1; LTF; CAMP; GYG1; PLD1; ATP11B; ANXA3; CARD6; MCTP1; CYSTM1; SLC36A1; HK3; SERPINB1; SRPK1; MAPK14; VNN1; ABCA13; GRB10; SLC37A3; CLEC5A; BMX; ADAM9; ASPH; UGCG; LCN2; TCN1; MMP8; CSGALNACT2; CLEC4D; PLBD1; FGD4; IRAK3; DRAM1; CKAP4; CAMKK2; KLFV; ATP8B4; ITGAM; HP; HPR; WSB1; MPO; MAFG; RAB31; PSTPIP2; BPI; MMP9; RETN; MCEMP1; CEACAM8; CD177; PGLYRP1; FCAR; CLTCL1; RGL4; TSPO; SAMSN1; KCNE1; TRPM2.

## Acknowledgements

This work was supported by the NIHR BioResource, the NIHR Cambridge Biomedical Research Centre and the NIHR Cambridge Clinical Research Facility. FV plasma assays were performed by the NIHR Cambridge Biochemical Assay Laboratory. FV constructs were prepared and expressed by Peak Proteins. Neutrophil proteins were characterised at the Mass Spectrometry Facility at the University of Dundee and the QMRI flow cytometry and cell sorting facility. Sequencing was supported by Paul Coupland from the CRUK Cambridge Institute Genomics Core.

The work was funded by awards from NIHR to the NIHR BioResource and the NIHR Cambridge Biomedical Research Centre, Evelyn Trust, UKRI/NIHR funding through the UK Coronavirus Immunology Consortium (UK-CIC) and a CSO award (COV/DUN/20/01). KGCS holds a Wellcome Trust Investigator award. BG holds an award from the Aging Biology Foundation Europe to BG. RKG holds a Wellcome Senior Fellowship (WT108082AIA). PK is supported by the Australian and New Zealand Society of Nephrology and the Royal Australasian College of Physicians. SRW holds a Wellcome Trust Senior Clinical Fellowship (209220). ERW holds a Wellcome Clinical Training Fellowship award (108717/Z/15/Z). NM was supported by a DFG Research Fellowship. PFC is a Wellcome Trust Principal Research Fellow (212219/Z/18/Z), and a NIHR Senior Investigator, and receives support from the Medical Research Council (MRC) Mitochondrial Biology Unit, the MRC International Centre for Genomic Medicine in Neuromuscular Disease, the Leverhulme Trust, a MRC research grant, and an Alzheimer’s Society Project Grant. JAN holds a Wellcome Trust Senior Research Fellowship (215477/Z/19/Z).

## Authors contributions

Conceptualisation JB, JSP, JW, PAL, KGCS data collection JW, PK, PAL, FM, LB, LT, KS, NW data curation JW, PK, FM, LB, LT, NW, KS, BG, JCM formal analysis JW, PK, MDM, NW, BG, JCM funding acquisition JB, PFC, NK, PAL, KGCS investigation JW, PK, PAL, FM, LB, LT, MDM, FJC-N, KB, NM, NKW, ERW, RKG, MT, MPW, JAN, SRW, KGCS methodology and study design JW, PK, PAL, MDM, FJC-N, KB, NM, NKW, ERW, PFC, NK, SP,KS, NW, RKG, MT, MPW, SRW, WHO, BG, MK, KGCS, JSP, JRB writing – original draft JW, PK, JSP, MDM writing – review & editing JW, PK, PAL, FM, LB, LT, RSA-L, MDM, FJC-N, KB, NM, NKW, ERW, PFC, NK, SP,KS, NW, RKG, MT, MPW, SRW, WHO, BG, MK, KGCS, JSP, JRB

